# High Incidence of Adverse Pregnancy Outcomes are Associated with Maternal Age and Infection Status in a Resource-Limited Community

**DOI:** 10.64898/2026.05.29.26354424

**Authors:** Sarah Naulikha Kituyi, Alfred Owino Odongo, Rachael Wachuka, Shadrack Wambua, Francis Kobia, Jesse Gitaka, Bernard N. Kanoi

## Abstract

Maternal health during pregnancy is critical for favorable birth outcomes and long-term wellbeing of both mothers and infants. Women in rural, malaria-endemic regions face unique biological and socioeconomic challenges that may increase the risk of adverse pregnancy outcomes (APOs). This study investigated the incidence and determinants of APOs among pregnant women attending antenatal care at Webuye sub-County Hospital in Western Kenya, a rural malaria-endemic setting.

We conducted a retrospective cohort analysis utilizing previously collected data of 300 women enrolled during early pregnancy and followed through delivery. Maternal demographic, clinical, and infection-related factors were assessed, and associations with APOs were evaluated using chi-square tests and multivariable logistic regression. Maternal age and gestational age at enrollment were significantly associated with malaria history (P<0.001). Maternal BMI abnormality (124.5/1000 pregnancies), anemia (99.3/1000), fetal or neonatal death (81.3/1000), and preterm birth (43.8/1000) were observed (all P<0.001), suggesting a substantial burden. Younger mothers (<20 years) and older mothers (>35 years) were significantly more likely to develop anemia (P =0.026), and prior malaria infection further increased anemia risk (P =0.02). Abnormal urinalysis findings indicative of urinary tract infection were significantly associated with low birthweight (P =0.031). No significant associations were found between APOs and infant sex, parity, gravidity, or maternal ABO blood type.

These findings highlight a substantial burden of APOs in this rural population, exceeding national and global estimates. Strengthening malaria prevention, nutritional support, urinary infection screening, and encouraging early antenatal care attendance are critical to improving maternal and neonatal outcomes. Targeted interventions for adolescent and older mothers, along with enhanced point-of-care diagnostics, may reduce preventable complications in similar resource-limited, malaria-endemic settings.

## INTRODUCTION

Maternal health during pregnancy is crucial for successful gestation and the survival of both the mother and the newborn. Biological, genetic, and socioeconomic factors significantly influence a woman’s health during and after pregnancy (1). These factors, in turn, affect pregnancy outcomes (2). Adverse pregnancy outcome (APO) collectively refers to the health related complications that may occur during pregnancy, delivery and after delivery to either the mother, the neonate, or infant (2,3). These complications include low birth weights (LBW), still births, various obstetric complications, neonatal (4), and maternal mortality (1).

In Kenya, the presentation of these APOs is evidently high (5) and the maternal mortality is estimated to be at 530 deaths for every 100,000 live births (6). It is estimated that about 12% of the births in Kenya are small for gestational age while 1.9% of the recorded deliveries are still births (5). The burden of LBW in Kenya is about 13%, with rural regions exhibiting the highest rates (7). The major causes of deaths among pregnant mothers in Kenya include unsafe abortions, sepsis, and hemorrhage and often exacerbated by limited access to skilled birth attendance (8). Perinatal mortality is largely attributed to low birth weight, preterm delivery, and inadequate perinatal care (9). These APOs contribute to long-term developmental challenges, including growth restriction and chronic cardiopulmonary disorders (5,10).

A recent study in Webuye sub-County, Western Kenya, reported no significant association between malaria in pregnancy and LBW (11) which might be attributed to enhanced intermittent malaria prevention using Sulfadoxine-Pyrimethamine (SP) (12) known to safeguard against LBW(13) by initiating gestational weight gain (14). The same study did not find significant associations between maternal anemia or diabetes and LBW. Building on these findings, and using a larger cohort, we aimed to examine additional maternal factors, including age, gravidity, parity, ABO blood type, and urinalysis results, not previously explored in the same population.

Comprehending the role of maternal factors in shaping pregnancy outcomes is very critical for birth preparedness and emergency obstetric care planning (15). Insights from such research can support early identification and management of high-risk pregnancies and inform public-health strategies to improve maternal and neonatal survival. Furthermore, identifying biological and clinical predictors of APOs may empower women and healthcare providers to make informed reproductive and maternal health decisions. This study therefore addresses an important knowledge gap in a rural, resource-limited setting, to investigate the incidence and determinants of APOs. Findings will support maternal health interventions for rural populations in Bungoma County and contribute to Sustainable Development Goal 3 (SDG-3) on improving maternal and newborn health.

## MATERIALS AND METHODS

### Study Design and Setting

This was a retrospective cohort analysis utilizing previously collected data from a prospective cohort study conducted at Webuye sub-county hospital by the Centre for Malaria Elimination, Mount Kenya University in March to December of 2022. Part of the initial study has been previously described (11). Briefly, the study hospital is in Webuye sub-county (code 3911), Bungoma County, Western Kenya. At the time of the data collection, the county had a population of 1,919,490 people where the estimated number of male individuals was 939,105 with an equal proportion of women estimated at 980,385 including 429,762 women of childbearing age (15-49 years) (16). The anonymized data was accessed in June to August of 2025 and the authors did not have any information that could identify the individuals in the study.

### Study Population and Data Collection

Pregnant mothers (aged between 18 and 45 years) attending the antenatal clinic at the Webuye sub-County Hospital were enrolled into the initial study between March and December 2022 and followed up until delivery (11). The current study assessed clinical and sociodemographic parameters including gestational age, maternal age, parity, gravidity, height, weight, and baby’s sex. Maternal outcomes assessed included newborn birthweight and survival, maternal blood group, anemia status, urinalysis results, and malaria history confirmed by microscopy.

### Inclusion and Exclusion Criteria

Eligible participants were pregnant women ≤16 weeks’ gestation at enrollment who had resided in Webuye sub-County for ≥6 months and provided written informed consent. Participants planning to relocate during the study period were excluded, as were women with known HIV status or severe medical conditions requiring referral outside the hospital at enrollment.

### Sample Size Determination

This study employed a census approach, including all 300 pregnant women enrolled at the facility in 2022. A prior study using a similar design demonstrated adequate statistical power with smaller groups (n = 60 per arm) (14). Therefore, inclusion of all eligible participants in the cohort was expected to yield ~80% power while minimizing sampling error.

### Data Analysis

Data were entered into Microsoft Excel and analysed using R version 4.4.1. Descriptive statistics summarized sociodemographic variables. Incidence rates of adverse pregnancy outcomes (APOs) were calculated as events per 1,000 valid pregnancies. A Chi-square goodness-of-fit test compared observed versus expected APO frequencies. Associations between maternal factors and APOs were evaluated using Chi-square or Fisher’s exact tests as appropriate. Multivariable logistic regression models adjusted for potential confounders including maternal age and malaria history were used as appropriate. Results are presented as odds ratios (ORs) with 95% confidence intervals (CIs). Statistical significance was set at P < 0.05.

### Ethical Approval

This study was approved by the Mount Kenya University ethical and scientific review committee and given the following ethical approval numbers: 1099 (MKU/ERC/2026) and 3841 (MKU/ISERC/5119) and was conducted as per the standards of the declaration of Helsinki. Additional approval for the parent study (MKU/ERC/2026) was granted by the National Commission of Science and Technology (NACOSTI) (Permit NACOSTI/P/22/22502). All the study participants confirmed their willingness to take part in the study and subsequently gave informed written consent. All participants data was anonymized and held in confidence.

## RESULTS

The sociodemographic characteristics of the current study population are described in Table 1. While a total of 300 individuals were enrolled, the summaries for the outcomes of interest are reported using varying sample sizes (valid cases; N) because available data differed across outcomes. This approach, known as available-case analysis (17), resulted in varying valid-case sample sizes across outcomes, a common phenomenon in real-world data settings due to incomplete entries and exclusion of cases with data-entry errors. To maintain analytic integrity, only observations with complete and valid outcome data were included, avoiding potential bias from imputation or incomplete records.

**Table 1:**
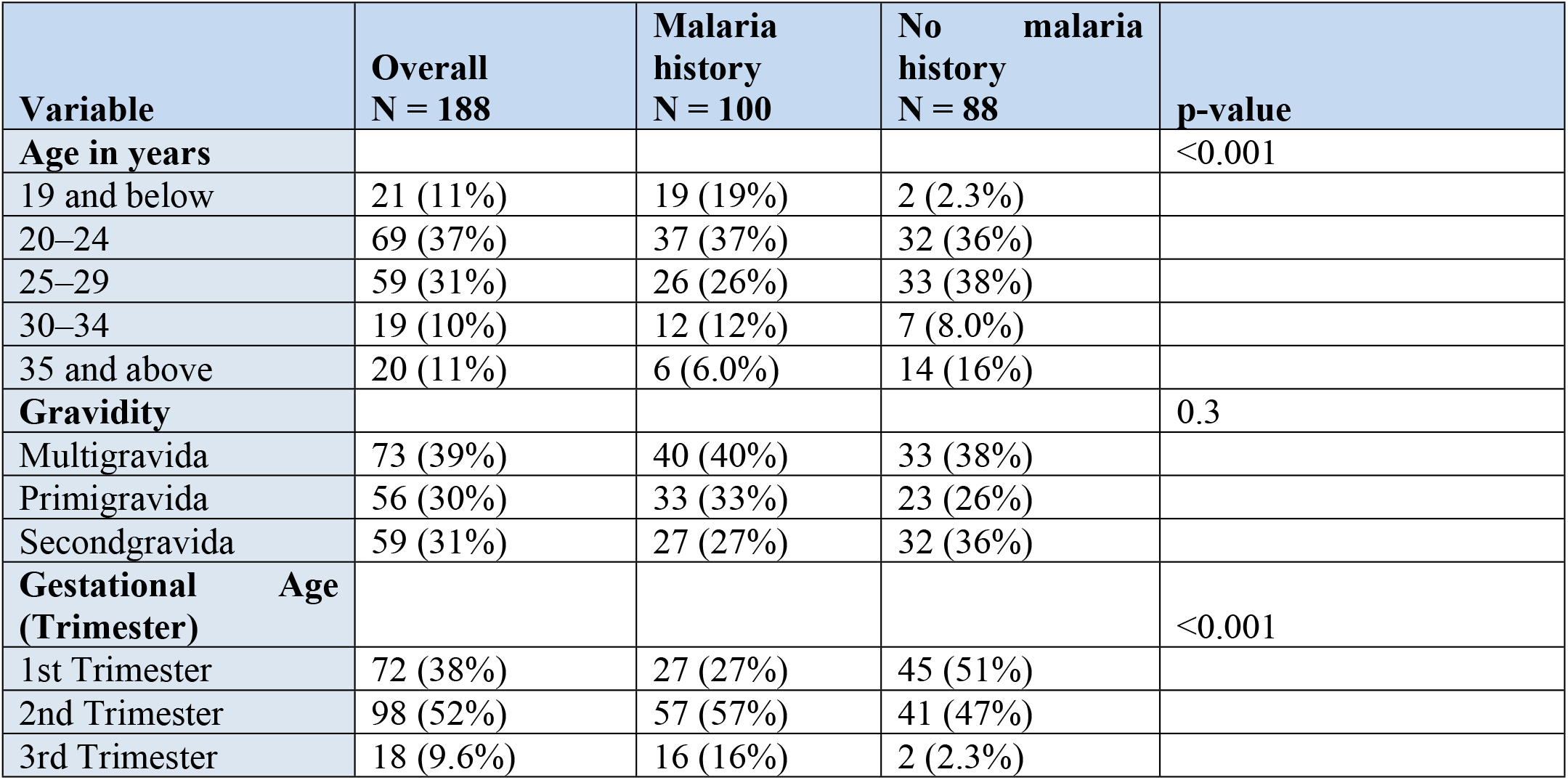
Summary of the Social Demographic Characteristics of the Study Populations.

### Social Demographic Characteristics

In Table 1, data was stratified by history of malaria infection given the high malaria burden in Western Kenya. This stratification was applied as malaria is highly prevalent in this region and may confound associations with pregnancy outcomes, consistent with Mukala et al., 2024.

The APOs evaluated included low birth weight, fetal and neonatal deaths, maternal anemia, maternal weight abnormalities and preterm births (Table 2). Key maternal factors examined included age, fetal sex, blood group, infection history, parity, and gravidity (18–21)(22,23) (24,25)(5)(26).

**Table 2:** Description of Adverse Pregnancy Outcomes.

| Variable | Description | Adverse Outcome Criteria |
| --- | --- | --- |
| Low birth weight | Baby's weight in grams | < 2.5kgs |
| Fetal and neonatal deaths | Newborn outcome | All cases of infant death, macerated, and fresh stillbirth, abortions, and miscarriages |
| Maternal anemia | Maternal hemoglobin | < 10.0 g/dL |
| Maternal BMI abnormality | Underweight, normal, overweight and obese based on maternal_bmi (derived from Weight and Height) | Underweight (<18.5), normal weight (18.5-24.9) overweight (25.0-29.9) and obese ($\geq 30.0$) |
| Preterm births | Weeks of gestation at delivery | < 37 weeks |

Low birth weight was defined as a birth weight of <2500 g (27). Fetal/neonatal deaths included infant deaths, still births, abortions and miscarriages. Maternal anemia was indicated by hemoglobin levels of <10.0 g/dL. Maternal BMI abnormality included underweight (<18.5) or obese (≥30.0). Preterm birth refers to delivery before 37 weeks of gestation (28).

### The Incidences of Adverse Pregnancy Outcomes

Maternal BMI abnormality (124.5/1000) (where the most frequent abnormality was overweight followed by obesity) and anemia (99.3/1000) were the most frequent APOs, followed by fetal/neonatal death (81.3/1000), preterm birth (43.8/1000), and low birth weight (33.6/1000) (Table 3). The Pearson Chi-Square tests indicated that all observed APO rates significantly differed from expected distributions (all P < 0.001), suggesting that these outcomes occurred at non-random, clinically relevant frequencies in the population. Further analysis showed a highly significant association between BMI category and adverse outcomes (χ^2^ = 96.912, df = 2, p < 0.001). Both underweight and obese participants had the highest risk, with 100% experiencing adverse outcomes, while normal/overweight participants had a much lower risk (16.3%). This suggests that extremes of BMI are strongly associated with adverse outcomes in this population.

**Table 3:** Incidence Rates of Key Adverse Pregnancy Outcomes.

| Adverse Outcome | Valid Cases (N) | Percent of Total (%) | Proportion with adverse Outcome (%) | Pearson Chi-Square | df | p-value | Incidence per 1,000 (=count/total x1000) |
| --- | --- | --- | --- | --- | --- | --- | --- |
| Low birth weight (<2.5 kg) | 268 | 88.7 | 3.4 | 233.209 | 1 | <0.001 | 33.58 |
| Fetal and neonatal deaths | 283 | 93.7 | 8.1 | 198.477 | 1 | <0.001 | 81.27 |
| Maternal anemia | 282 | 93.4 | 9.9 | 181.121 | 1 | <0.001 | 99.29 |
| Preterm birth (<37 weeks) | 274 | 90.7 | 4.4 | 228.102 | 1 | <0.001 | 43.79 |
| Maternal BMI abnormality | 273 | 90.4 | 12.5 | 153.938 | 1 | <0.001 | 124.54 |

### Association of Maternal Age and Infant Sex with APOs

The association between maternal age and APOs was examined using Chi-square or Fisher’s exact tests. Mothers aged ≤20 or >35 years had a significantly higher prevalence of anemia compared with those aged 21–35 years (χ^2^(1, N = 283) = 4.969, P = 0.026; 16.7% vs. 7.6%) (**Figure 1**). No significant associations were observed between maternal age and other APOs (**Table 4**), nor between infant sex and any APO.

**Table 4:** The Association Between the Maternal Age, the Sex of the Baby and the Occurrence of the other Adverse Pregnancy Outcomes.

| Adverse Outcome | Grouping Variable | Valid Cases (N) | Cases with Outcome (n) | Proportion (%) | Test Used | Test Statistic | df or Method | p-value |
| --- | --- | --- | --- | --- | --- | --- | --- | --- |
| Low birth weight (<2.5 kg) | maternal_age_group | 268 | 9 | 3.4 | Fisher's Exact |  | Exact | 0.46 |
| Fetal& neonatal deaths (e.g. stillbirth) | maternal_age_group | 262 | 4 | 1.5 | Fisher's Exact |  | Exact | 0.641 |
| Preterm birth (<37 weeks) | maternal_age_group | 274 | 12 | 4.4 | Fisher's Exact |  | Exact | 0.185 |
| Maternal BMI abnormality (underweight or obese) | maternal_age_group | 273 | 34 | 12.5 | Chi-Square | 1.583 | 1 | 0.208 |
| Low birth weight (<2.5 kg) | sex_group | 266 | 8 | 3 | Fisher's Exact |  | Exact | 0.34 |
| Fetal& neonatal deaths (e.g. stillbirth) | sex_group | 262 | 3 | 1.1 | Fisher's Exact |  | Exact | 0.601 |
| Maternal anemia | sex_group | 248 | 27 | 10.9 | Fisher's Exact |  | Exact | 0.751 |
| Preterm birth (<37 weeks) | sex_group | 264 | 4 | 1.5 | Fisher's Exact |  | Exact | 0.353 |
| Maternal BMI abnormality (underweight or obese) | sex_group | 239 | 29 | 12.1 | Fisher's Exact |  | Exact | 0.878 |

**Figure 1:**
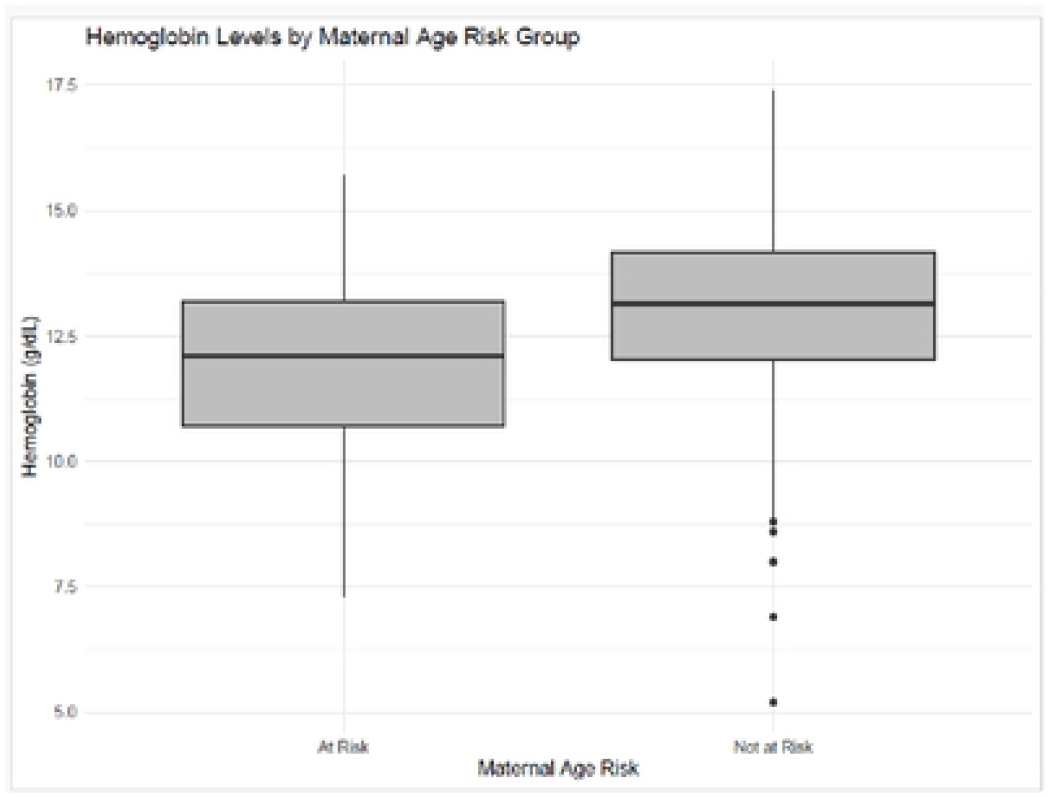
Box plot illustrating the association between maternal age and the risk of anemia

### Association of Gravidity and Parity with APOs

Cross-tabulation and Chi-square tests showed no statistically significant association between gravidity and APOs (χ^2^(6, N = 246) = 3.38, P = 0.760) or between parity and APOs (χ^2^(5, N = 246) = 4.75, P = 0.447) (**Tables 5 & 6**). Although not statistically significant, gravidity 4 exhibited the highest proportion of APOs (42.1%). A logistic regression adjusting for maternal age and malaria history also showed no significant association between gravidity or parity and APOs.

**Table 5:** Association between Gravidity and Adverse Outcomes.

| Gravidity | Total N (%) | No Adverse Outcome N (%) | Yes Adverse Outcome N (%) | % with Adverse Outcome | Pearson Chi-Square | df | P-value |
| --- | --- | --- | --- | --- | --- | --- | --- |
| 1 | 69(28.0%) | 44 (63.8%) | 25 (36.2%) | 36.2% | 3.375 | 6 | 0.760 |
| 2 | 67(27.2%) | 49 (73.1%) | 18 (26.9%) | 26.9% |  |  |  |
| 3 | 69(28.0%) | 50 (72.5%) | 19 (27.5%) | 27.5% |  |  |  |
| 4 | 19(7.7%) | 11 (57.9%) | 8 (42.1%) | 42.1% |  |  |  |
| 5 | 18(7.3%) | 12 (66.7%) | 6 (33.3%) | 33.3% |  |  |  |
| 6 | 3(1.2%) | 2 (66.7%) | 1 (33.3%) | 33.3% |  |  |  |
| 14 | 1(0.4%) | 1 (100%) | 0 (0.00%) | 0% |  |  |  |
| Total | 246(100%) | 169 (68.7%) | 77 (31.3%) | 31.3% |  |  |  |

**Table 6:** Association between Parity and Adverse Outcomes.

| Total Parity | No Adverse Outcome N (%) | Yes Adverse Outcome N (%) | Total | % of Total Adverse Outcomes | Pearson Chi-Square | df | P-value |
| --- | --- | --- | --- | --- | --- | --- | --- |
| 0 | 45 (65.2%) | 24 (34.8%) | 69 | 28.0% | 4.750 | 5 | 0.447 |
| 1 | 49 (71.0%) | 20 (29.0%) | 69 | 28.0% |  |  |  |
| 2 | 51 (75.0%) | 17 (25.0%) | 68 | 27.6% |  |  |  |
| 3 | 9 (50.0%) | 9 (50.0%) | 18 | 7.3% |  |  |  |
| 4 | 13 (68.4%) | 6 (31.6%) | 19 | 7.7% |  |  |  |
| 5 | 2 (66.7%) | 1 (33.3%) | 3 | 1.2% |  |  |  |
| Total | 169(68.7%) | 77 (31.4%) | 246 | 100.0% |  |  |  |

### Association Between Maternal Blood Group and Occurrence of APOs

Maternal blood group was not significantly associated with APOs (χ^2^(8, N = 246) = 3.516, P = 0.898; **Table 7**). Logistic regression adjusting for maternal age, gravidity, parity, and malaria history yielded a model approaching significance (χ^2^(18) = 27.64, P = .068), but no individual predictor reached P < .05.

**Table 7:** Distribution of Adverse Pregnancy Outcomes by Maternal Blood Group using Chi-Square Test of Association (N= 246).

| Maternal Blood Group | No Adverse Pregnancy Outcome | Adverse Pregnancy Outcome | Total (N) | % Within Adverse Outcome | Pearson Chi-Square | df | P-Value |
| --- | --- | --- | --- | --- | --- | --- | --- |
| AB- | 1 | 0 | 1 | 0% | 3.516 | 8 | 0.898 |
| AB+ | 5 | 2 | 7 | 0.8% |  |  |  |
| A- | 1 | 0 | 1 | 0% |  |  |  |
| A+ | 43 | 20 | 63 | 8.1% |  |  |  |
| B- | 2 | 0 | 2 | 0% |  |  |  |
| B+ | 41 | 18 | 59 | 7.3% |  |  |  |
| O- | 2 | 0 | 2 | 0% |  |  |  |
| O+ | 73 | 37 | 110 | 15.0% |  |  |  |

### Associations Between Maternal Infections and the Occurrence of APOs

History of malaria was significantly associated with maternal anemia (P = 0.02). Abnormal urinalysis was associated with LBW (P = 0.032, **Figures 2 and 3**). Gestational age at enrollment was associated with anemia (P = 0.003), with highest burden in second-trimester enrollees (**Table 8**). No other APOs were associated with gestational age

**Table 8:** The association between the gestation age at enrollment and anemia in pregnancy.

| Adverse Outcome | Gestation trimester | N | Adverse Outcome Presence | Proportion | Test Used | df or Method | p-value |
| --- | --- | --- | --- | --- | --- | --- | --- |
| Maternal anemia | First Trimester | 119 | 4 | 3.4 | Fisher's Exact | Exact | 0.00286 |
| Maternal anemia | Second Trimester | 142 | 22 | 15.5 | Fisher's Exact | Exact |  |
| Maternal anemia | Third Trimester | 21 | 2 | 9.5 | Fisher's Exact | Exact |  |

**Figure 2.**
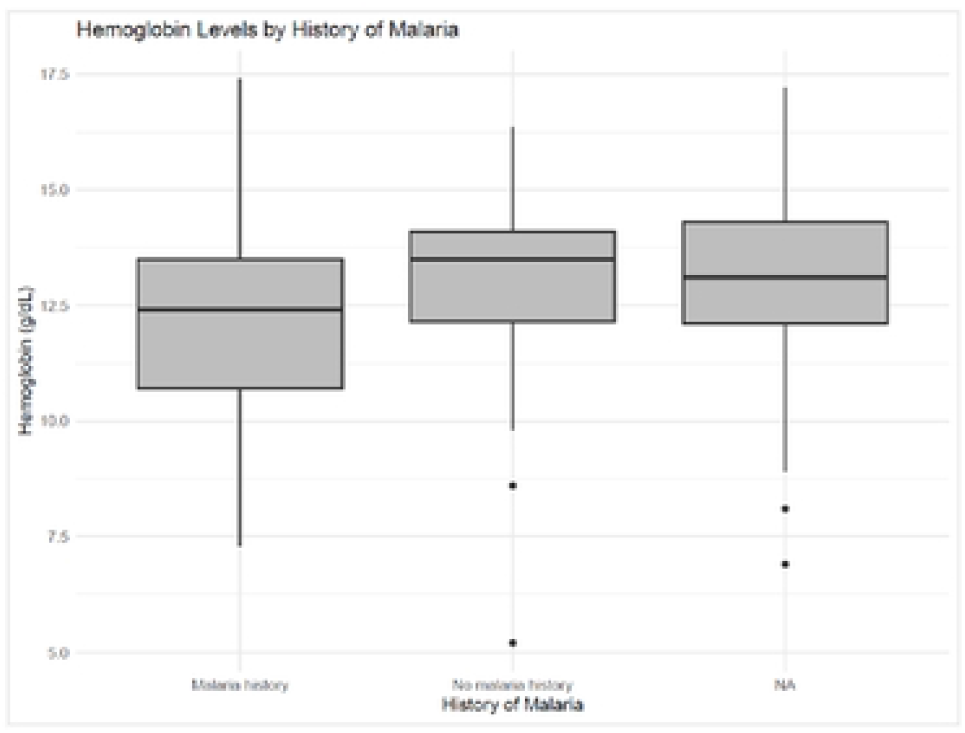
A box plot illustrating the association between the history of malaria and anemia (defined by hemoglobin levels).

**Figure 3.**
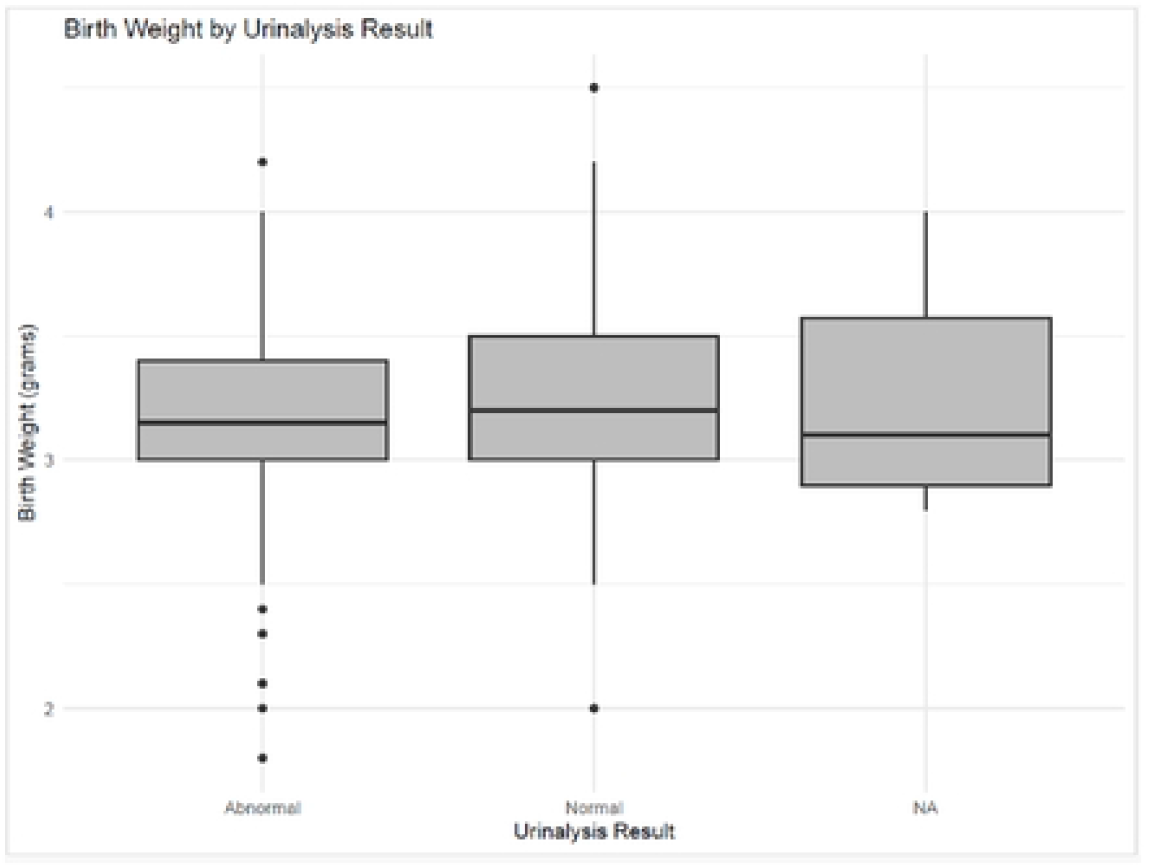
Box plot of the analysis between urinalysis results and low birth weights.

## DISCUSSION

This study investigated the incidence of APOs and associated factors among pregnant women in Webuye sub-County, a malaria endemic region. Sociodemographic data were stratified by malaria history to identify groups at higher risk of APOs in whom malaria may confound outcomes. There was a statistically significant relationship between age group and malaria status during pregnancy (P<0.001), where relatively younger women (especially <24 years) had a higher proportion of malaria cases, suggesting lower immunity or reduced prior exposure (29). Similarly, women in the first and second trimester were more likely to have malaria than those in the third trimester (P < 0.001), consistent with trends reported by Takem & D’Alessandro, 2013. This pattern may reflect immature pregnancy-related immune adaptation in younger women and early pregnancy immunological vulnerability (29).

Overall, APO incidence in this rural setting was high, suggesting a disproportionate burden compared to national estimates. The incidence of maternal anemia exceeded national levels (~40%) (30,31) and surpassed the WHO 2025 target of <50% (32), indicating a substantial burden of nutritional deficiency and malaria-associated anemia (11). Fetal and neonatal mortality rates were also higher than Kenyan estimates (18–21/1,000 live births) (33), potentially hindering progress toward SDG targets of ≤12 deaths/1,000 (33). Preterm birth incidence exceeded global estimates (~10%) (34), pointing to gaps in antenatal and obstetric care in this rural population. Although LBW rates were lower than those reported in Kilifi (35), they remain above WHO 2025 reduction targets (35). Collectively, these findings reflect structural health inequities, infectious disease burden, and nutritional vulnerability in this setting.

Maternal anemia was significantly associated with age. Younger mothers may experience anemia due to competition for nutrients during ongoing physiological growth (21), while older mothers (>35 years) may face nutrient depletion due to repeated pregnancies and higher comorbidity burden (36). Mothers aged 21–35 likely benefit from physiological maturity and more stable health-seeking behavior (28). Contrary to studies suggesting sex-specific neonatal vulnerability(23), fetal sex was not associated with APOs, potentially due to differing outcome definitions or confounding factors such as maternal age and infection burden (37).

Although gravidity four showed the highest proportion of APOs, followed by primigravidity, no statistical significance was observed. Primigravidas often face higher APO risk due to immunological naivety and limited ANC experience (38), while high-gravidity mothers may experience uterine vascular remodeling and nutrient depletion (39). The lack of statistical significance here may reflect sample size limitations and residual confounding. Parity similarly showed no association with APOs.

Maternal blood group was not associated with the occurrence of any APOs. It is worth noting that many of the maternal blood groups (like AB-, A-, B-, and O-) had very small sample sizes, each with only 1 to 2 participants), which reduced the statistical power of the test. However, existing evidence in literature equally suggests inconsistent associations between the maternal blood group and APOs (40). A recently study by Rom et al., 2024 did not find any significant association between the maternal ABO blood and APOs consistent with our findings.

Consistent with Mukala et al., 2024, malaria history was not associated with LBW but was associated with maternal anemia. Mechanistically, *P. falciparum* infects and destroys red blood cells (41)(42), induces inflammatory cytokines that suppress erythropoiesis (43,44), alters iron metabolism (45), and may cause placental sequestration leading to impaired nutrient transport (11)(46). Pregnancy increases iron demand (47), compounding susceptibility. The observed lack of a statistically significant association between the history of malaria and LBW in this study population may be driven by the reported use of SP for intermittent malaria prevention in pregnancy (12) which is expected to safeguard against LBW(13).

Abnormal urinalysis findings were associated with LBW. Presence of pus cells suggests undiagnosed urinary tract infection and inflammation, which can impair placental function and fetal growth (48)(49)(50). Yeast cells may indicate candidiasis-related dysbiosis and immune activation, which has been linked to APOs (51). These findings highlight undiagnosed maternal infections as modifiable risk factors and signal the need for improved routine ANC screening. Future research should incorporate nutritional status, hygiene practices, and antibiotic use to further elucidate these relationships.

A major limitation is that the cohort was drawn only from women attending routine ANC, meaning expectant mothers who utilize traditional midwives outside the health system were not represented, potentially leading to an underestimation of the true problem’s magnitude in the community. Therefore, larger longitudinal studies are warranted to clarify causal pathways and inform more robust, targeted interventions.

In conclusion, in this malaria-endemic rural population, adverse pregnancy outcomes were associated with a combination of infectious, nutritional, and maternal factors. Prior malaria infection increased the likelihood of maternal anemia, while abnormal urinalysis suggestive of underlying infections was strongly linked to LBW. Younger and older mothers and those enrolled early in pregnancy showed greater vulnerability to certain outcomes. These findings underscore the urgent need to strengthen early ANC attendance, enhance malaria prevention and treatment, and expand routine screening for anemia and maternal infections. The study strongly suggests that improving maternal nutrition and bolstering the diagnostic capacity at primary facilities for rapid infection screening are crucial steps to reduce preventable complications in this and similar resource-limited settings.

## Data Availability

All data depicted in the manuscript is readily available on request from our repository at the center for malaria elimination-institute of tropical medicine-Mount Kenya University, Thika, Kenya.

## Abbreviations

APOs: Adverse pregnancy outcomes
WHO: World health organization
BMI: Basal metabolic index
LBW: Low birth weight
KDHS: Kenya Demographic and Health Survey
SDG: Sustainable development goal

## Acknowledgements

We appreciate the study volunteers from Webuye Level V Hospital, Bungoma county; and thank the research teams at Centre for Malaria Elimination, Mount Kenya University, for their technical assistance in processing the field samples and data.

